# Detection of pneumococcal carriage in asymptomatic healthcare workers

**DOI:** 10.1101/2024.07.19.24309369

**Authors:** Pari Waghela, Raechel Davis, Melissa Campbell, Rupak Datta, Maikel S. Hislop, Noel J. Vega, Loren Wurst, Devyn Yolda-Carr, Luke Couch, Michael Hernandez, Lindsay R. Grant, Ronika Alexander-Parrish, Adriano Arguedas, Bradford D. Gessner, Richard A. Martinello, Daniel M. Weinberger, Anne L. Wyllie

## Abstract

**Background:** Healthcare workers are at increased risk of exposure to respiratory pathogens including *Streptococcus pneumoniae* (pneumococcus). While little asymptomatic carriage has been reported in young-to-middle-aged adults, this may be due to non-sensitive diagnostic methods. The aim of the study was to investigate the rates of pneumococcal carriage in a large cohort of healthcare workers using saliva as a respiratory specimen.

**Methods:** We evaluated the prevalence of pneumococcal carriage in a convenience sample of saliva, self-collected from asymptomatic healthcare workers at Yale New Haven Hospital (CT, USA) who were testing for SARS-CoV-2 from March 30 to June 11, 2020. Samples were transported at ambient temperature and stored at −80°C within 12 hours. DNA extracted from the culture-enriched saliva was later tested using qPCR for *piaB, lytA*, and serotype. Saliva samples were considered positive for pneumococcus when the *piaB* Ct value was <40.

**Results:** Study participants were 22-74 years old (mean=38.5), 75% female, 75% white, with occupations including registered nurses (48%), medical doctors (23%), and patient care assistants (5%). Overall, 138/1241 (11%) samples from 86/392 (21%) individuals tested *piaB*-positive for pneumococcus at some point during the 4-month study period, with 28 (33%) colonized individuals positive at multiple time points. Carriers reflected the overall study population. No significant demographic characteristics were associated with detection of pneumococcus. Colonized individuals primarily carried serotypes 19F (25%) and 3 (12%), however, we were unable to resolve a primary serotype for 31% of all pneumococcus-positive samples identified.

**Conclusions:** During a period of mandatory masking, we identified a cumulative pneumococcal carriage prevalence of 21% among healthcare workers. This study highlights that healthcare workers may act as unrecognized reservoirs of pneumococcus in the population. Despite long-standing PCV7 and PCV13 pediatric immunization programs, vaccine serotypes continue to be prevalent among the adult population.

## INTRODUCTION

A common commensal inhabiting the upper respiratory tract, *Streptococcus pneumoniae* (pneumococcus) is also a leading cause of respiratory infections such as otitis media and sinusitis and life-threatening, invasive pneumococcal diseases (IPDs) including pneumonia, bacteremia, and meningitis. Children are considered the reservoir of pneumococcus in the population, with carriage rates typically reported to decline with increasing age, with little asymptomatic carriage reported in young-to-middle-aged adults.^1^ Prior studies have demonstrated that contact with children is associated with higher pneumococcal carriage prevalence in adults,^1,2^ including healthcare workers working in the pediatric setting.^3^ Since healthcare workers are often exposed to the respiratory secretions of sick individuals, it is not just those working in pediatrics who are potentially at an increased risk of exposure to respiratory pathogens including pneumococcus.

Pneumococcal carriage has seldom been investigated and reported among healthcare workers, despite their potentially elevated risk. Elucidating the prevalence of pneumococcal carriage in this population is not only important for informing occupational risks, but for understanding potential transmission dynamics within healthcare settings. Healthcare workers who are frequent carriers of pneumococci may pose an increased risk of transmission to vulnerable populations such as individuals who are elderly, immunocompromised, or have chronic illnesses. Understanding this risk can inform the development of strategies to minimize spread among both healthcare workers and patients. Therefore, in the current study, we evaluated pneumococcal carriage rates and serotypes in a cohort of healthcare workers at Yale New Haven Hospital.

## METHODS

### Ethics

Medical staff at Yale New Haven Hospital were eligible to participate in the study voluntarily. All study participants were enrolled and sampled in accordance with the Yale University Human Investigation Committee-approved protocol #2000027690. Demographics, clinical data, and samples were collected after the study participant acknowledged that they understood the study protocol and provided informed consent. All participant information and samples were collected in association with non-individually identifiable study identifiers.

### Participant enrollment

Between March 30 and June 11, 2020, healthcare workers without fever or respiratory symptoms working within Yale New Haven Hospital, on inpatient COVID-19 and non-COVID-19 units and emergency departments and ambulatory or with other occupational exposure to patients, were invited to enroll in the Yale IMPACT biorepository study.^4^ This study was designed to actively monitor asymptomatic healthcare workers for SARS-CoV-2 infection. Individuals were not included in the study if they were non-English speaking or under 18 years of age.

### Sample collection and processing

Healthcare workers were invited to self-collect saliva every three days for up to 84 days, or until testing positive for SARS-CoV-2. Samples were stored at +4°C until transport to the research laboratory where they were tested for SARS-CoV-2.^4^ The remnant sample volume was stored at −80°C within 12 hours. Starting July 2021, saliva samples were thawed on ice, vortexed to resuspend, and 100 μl were plated on TSAII plates with 5% sheep’s blood and 10% gentamicin, as previously described.^2^ Following overnight incubation, bacterial growth was harvested into brain heart infusion (BHI) medium supplemented with 10% glycerol. These culture-enriched saliva samples were then stored at −80 °C until further analysis.

### Pneumococcal detection

Culture-enriched samples were thawed on ice, then DNA was extracted from 200 μl of each sample using the MagMAX Viral/Pathogen Nucleic Acid Isolation Kit on the KingFisher Apex using a modified protocol.^2^ Each DNA template was tested using qPCR targeting pneumococcal-specific genes *piaB* and lytA, as previously described.^2^ Samples were considered positive with *piaB* Ct values <40.

### Strain isolation

Saliva samples that tested positive for *piaB* with a Ct <28 were re-visited by culture^2^ or magnetic bead-based separation (MBS)^5^ in an attempt to isolate pure pneumococcus. Isolates were serotyped by latex agglutination.

### Serotype determination

DNA extracts that tested positive for *piaB* were pooled by 4, those that tested negative were pooled by 10. Where possible, samples from the same individual were pooled together. Each pool was tested in 8 multiplexed serotyping assays^6^ targeting a total of 39 serotypes (see Supplementary Information).^7–10^ From each pool, 8 μl of DNA was tested in a total reaction volume of 25 μl. All samples from any pool generating a serotype-specific Ct value <40 were tested individually in that serotyping assay. The primary serotype of an individual sample was determined based on concordance between the *piaB* Ct value and the serotype-specific Ct value. Assay reliability was determined based on the lack of serotype-specific signal in *piaB*-negative pools or of serotype-specific signal >4 Ct lower than the *piaB* Ct value of positive samples.

### Data analysis

Differences in characteristics between pneumococcal carriage groups were tested using the Kruskal–Wallis rank sum test (continuous variable) or Fisher’s exact test (categorical variable). Estimates were considered statistically significant at *p*<0.05. Graphical visualizations and statistical analyses were performed in RStudio v.2022.07.2+576, using R v.4.1.3.^11^ Data preprocessing and visualization was performed using tidyverse^12^ and table1^13^ R packages.

## RESULTS

### Study population

A total of 525 healthcare workers provided informed consent for inclusion in the Yale IMPACT biorepository study.^4^ Of those, 16/525 (3%) tested positive for SARS-CoV-2^14^ and were excluded from this study due to limited power to describe co-infection. Saliva samples from 483/525 (92%) study participants were available for further analyses. From this cohort, we selected 392/483 (81%) study participants to test for pneumococcus. Study participants were selected to represent the demographic makeup of the total study population. Full demographic characteristics of the study population are shown in Table 1. Vaccination status, medical history nor living situation of study participants was available.

**Table 1.**
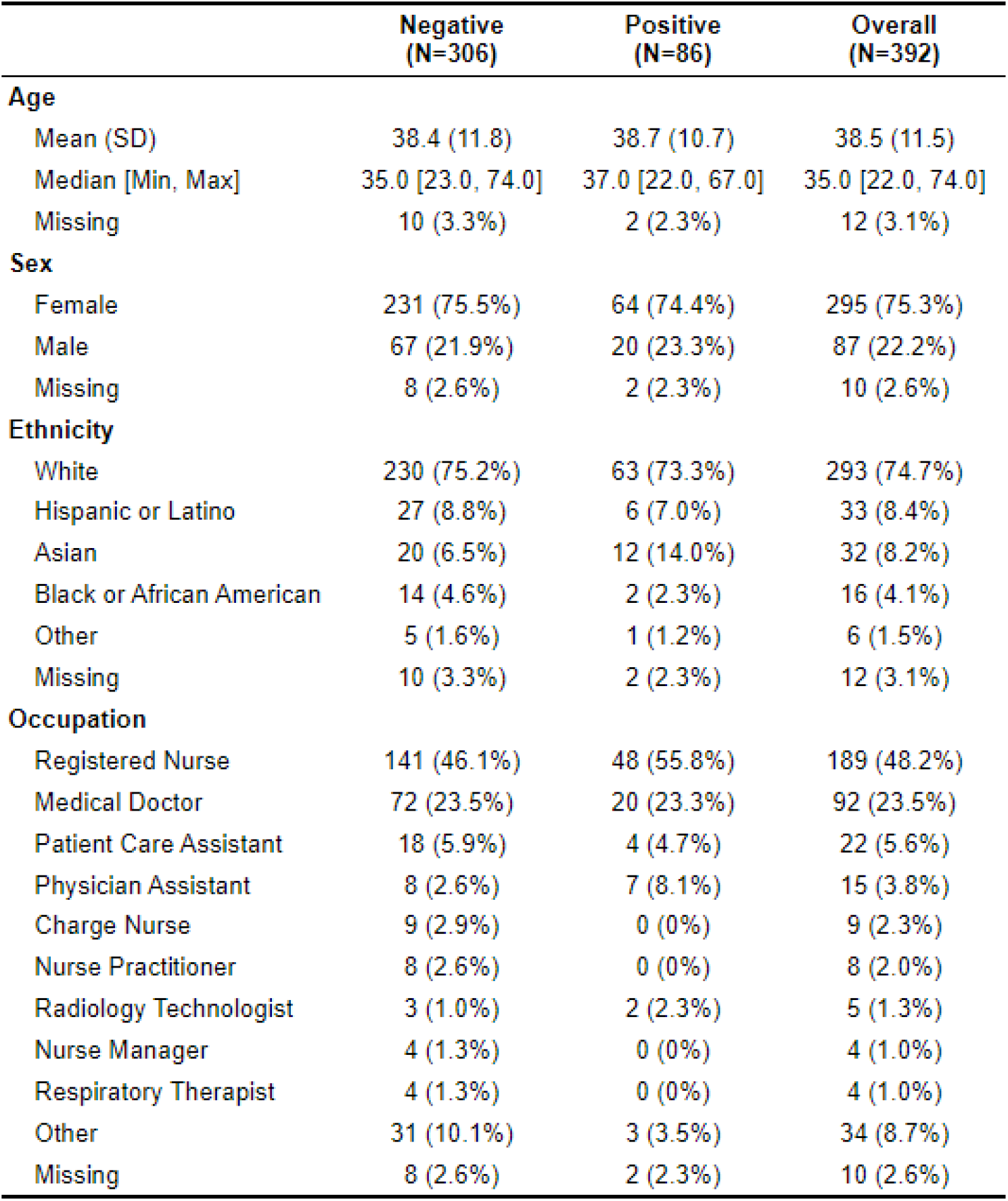
Characteristics of the study population, overall and by detection of pneumococcal carriage.

### Pneumococcal carriage

Overall, 1241 samples from 392 individuals (range=1-10 samples/individual, mean=3.17 samples/individual) were tested for pneumococcus. Of these, 138/1241 (11%) samples tested qPCR-positive for *piaB*, with 86/392 (21%) testing positive on at least one sampling moment during the 4-month study period (Figure 1); 28/86 (33%) carriers tested positive more than once (Figure 1). The average number of days between positive sampling moments was 15.2 days, while the shortest number of days between positive samples was 4 days and the longest 53 days.

**Figure 1.**
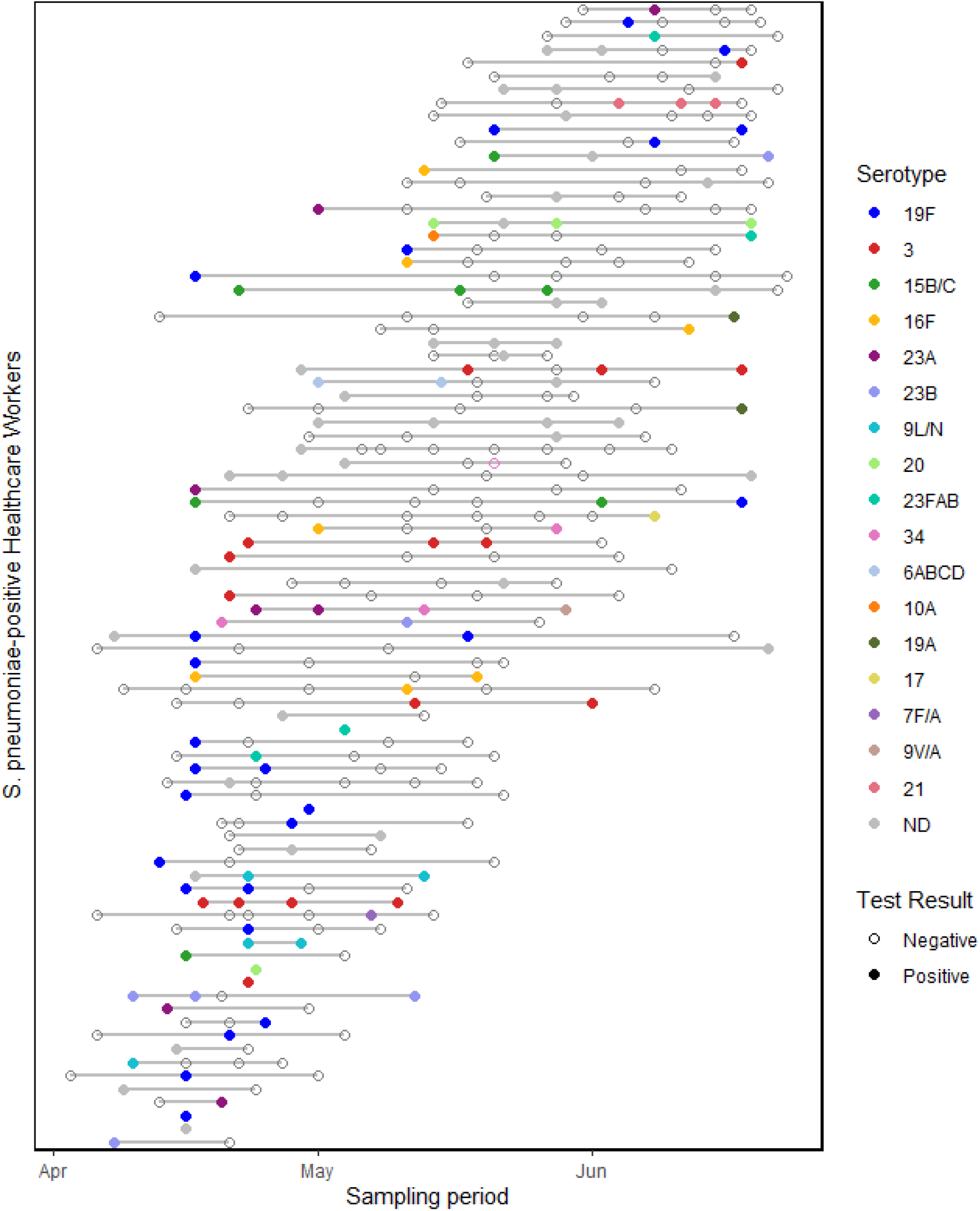
Detection of pneumococcus and pneumococcal serotype in self-collected saliva samples from asymptomatic healthcare workers, April-July 2020. All samples from any individual who tested qPCR-positive for pneumococcus (*piaB*) at least once during the study period are shown. Consecutive samples from the same individual are joined by a solid gray line. Solid circles denote samples qPCR-positive for *piaB*, colored according to primary serotype result. Secondary serotypes detected are not shown. Gray solid circles (“ND”) represent samples for which a serotype could not be determined using the available qPCR assays. Open circles denote samples qPCR-negative for *piaB*. The serotypes in the legend are ordered by decreasing prevalence among the study population.

Positive individuals reflected the overall study population (Table 1) and were 22-67 years old (mean=38.7 years), 74% female, and 73% white, with occupations including registered nurse (55%), medical doctor (23%), physician assistant (8%), and patient care assistant (4%). There were no significant demographic characteristics associated with *piaB*-positivity.

Positivity for pneumococcal carriage was defined as detection of *piaB*^2^ due to known non-specificity of the *lytA* qPCR assay leading to cross-detection of other Streptococci.^15^ In line with this, 287/1231 (23%) samples from 145/392 (37%) individuals tested positive for *lytA* (Ct <40) but negative for *piaB* (Ct >40). In contrast, 18 (1.5%) samples from 15 (4%) individuals were positive for *piaB* but negative for *lytA*. These *piaB*-positive/*lytA*-negative results were confirmed through re-testing. In addition, 8/18 (44%) of the *piaB*-positive only samples from 6/15 (40%) individuals also returned a serotype-specific Ct value that aligned with that obtained for *piaB*.

### Serotype determination

Of the 27 singleplex serotyping assays within the 8 multiplexed assays, we determined 20 of the assays to be reliable (2,^10^ 3,^10^ 6ABCD,^16^ 7BC/40,^7^ 7F/A,^10^ 9L/N,^10^ 9V/A,^7^ 10A,^10^ 11ADE,^10^ 14,^16^ 15B/C,^10^ 16F,^16^ 17F,^10^ 19A^8^ 19F,^10^ 21,^10^ 23A,^10^ 23B,^10^ 23FAB,^7^ 34^10^) representing 31 of 106 pneumococcal serotypes. Overall, 25% of pneumococcus-positive individuals (27 samples) tested positive for serotype 19F at least once over the study period and 12% of positive individuals (20 samples) tested positive at least once for serotype 3. Primary serotypes detected are presented in Figure 1., and all serotypes detected within all pneumococcus-positive individuals are presented in Figure 2; 7BC/40 was only detected as a secondary serotype, generating sinal in two samples from one individual 6 days apart.

**Figure 2.**
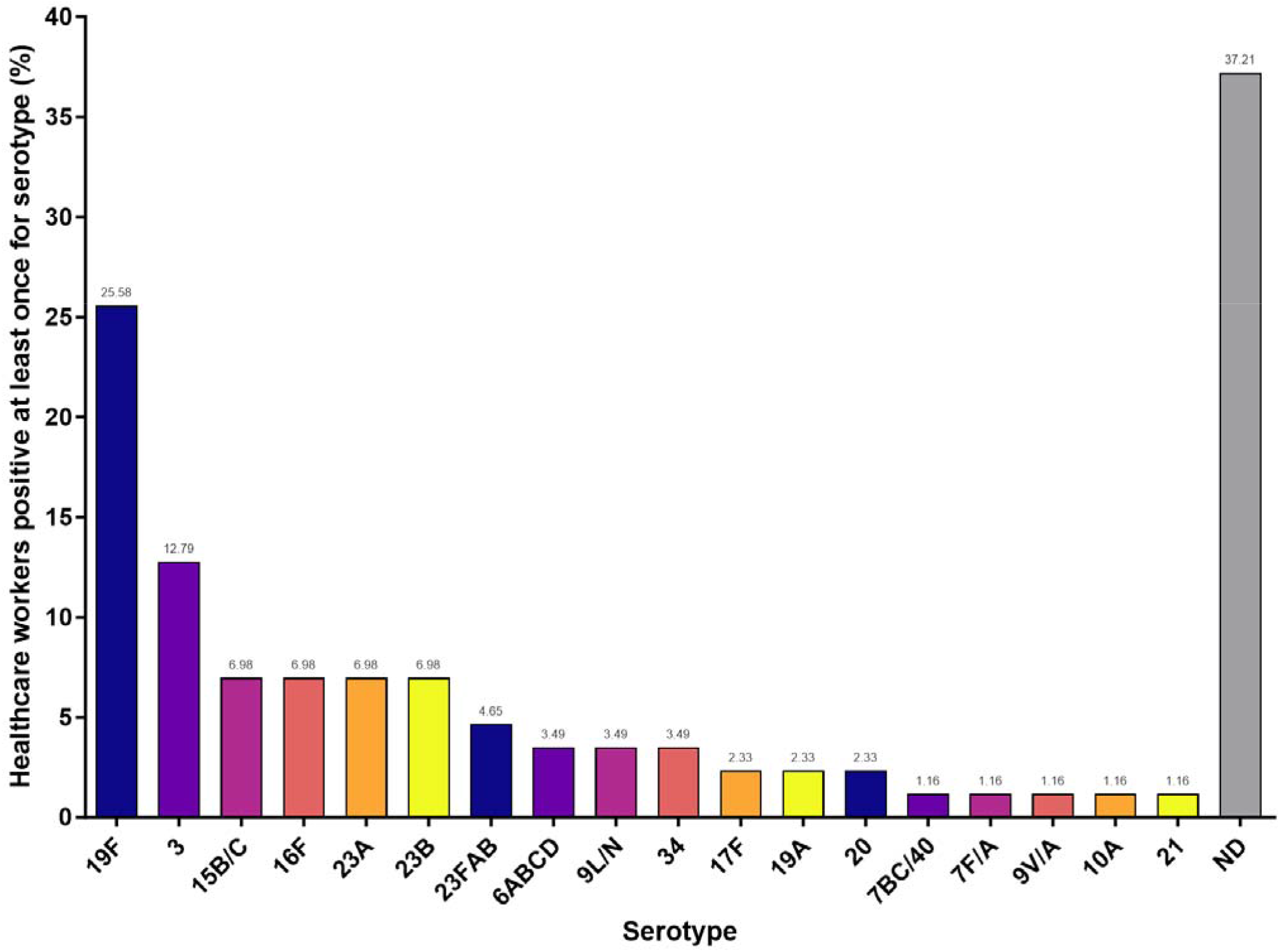
Percentage of healthcare workers who tested positive at least once for the pneumococcal serotypes detected in the study population. The percentage of positive individuals is also shown at the top of each bar. ND = a serotype could not be determined using the available assays.

Pneumococcal isolates were recovered from four samples. Three isolates from two individuals were confirmed to be serotype 3 and one isolate from another individual was confirmed to be serogroup 23 by latex agglutination.

## DISCUSSION

Most previous adult carriage studies have reported low pneumococcal prevalence^17,18^ based on nasopharyngeal swabs tested using conventional culture methods. We and others have demonstrated that testing oral samples using qPCR significantly increases the sensitivity of carriage detection in adults.^19–21^ Applying this approach to improve pneumococcal carriage detection among young-to-middle-aged adults also revealed that pneumococcal colonization is more prevalent among parents of young children as compared to adults without children.^1^ A similar finding was recently reported among healthcare workers of a pediatric hospital,^3^ likely also due to the high risk of transmission from children. In this study we hypothesized that healthcare workers in general might be at a greater risk of pneumococcal exposure. Our findings confirmed that the higher prevalence of pneumococcal carriage is not limited to those working with children but is more widespread across the entire hospital setting. The source of carriage acquisition, whether from the patient population or outside contacts, such as from children, is unclear.

The point prevalence of 11% and period prevalence of 21% for pneumococcal carriage in this study population was observed during the early COVID-19 pandemic when strict non-pharmaceutical interventions for mitigating SARS-CoV-2 transmission were in place and strictly adhered to, and IPD had declined globally. While we do not have data on the carriage prevalence amongst this population of healthcare workers prior to the COVID-19 pandemic, a number of studies, including our own study of older adults living in the same New Haven community,^2^ have demonstrated that pneumococcal carriage persisted in both children^22,23^ and adults^2^ during this time, despite the non-pharmaceutical interventions in place. It is therefore possible, that in the typical absence of these transmission mitigation measures and with a return to the typical annual circulation of respiratory pathogens^24^ and their associated respiratory infections, a higher rate of pneumococcal transmission in this population, and as such, higher rates of colonization may occur.

While the recommended sample type for the detection of pneumococcus in adults is the nasopharyngeal swab, with the addition of an oropharyngeal swab when possible,^25^ saliva has been effective for enhancing detection.^1,19,26^ Combined with qPCR, this is one of the most sensitive approaches for detecting pneumococcus.^27^ While in the current study, self-collected saliva samples were tested, in the recent study by Steurer et al. of pediatric healthcare workers,^3^ oropharyngeal swabs were tested using qPCR targeting *lytA* only. Despite its higher sensitivity, we have recently demonstrated non-specificity of the *lytA* assay when applied to saliva,^15^ due to the cross-detection of gene homologues present in non-pneumococcus Streptococci. Steurer *et al*. recognized this as a potential limitation of their study.^3^ While the specificity of results would need to be confirmed through testing for additional gene targets,^28^ the prevalence detected in pediatric healthcare workers is not too different to our own observations.

This risk of cross-detection also applies to molecular methods of serotype detection,^29^ with non-pneumococcal Streptococci also harboring capsular biosynthesis genes which can be at least, in part, identical to pneumococcal genes. We applied a strict assessment of the specificity of each qPCR assay through the testing of negative samples and critically evaluating the Ct values generated by assays of higher specificity to determine serotyping reliability. We observed a high prevalence of serotype 19F, detected at least once in 25% of carriers and of serotype 3, detected at least once in 12% of carriers, with the observed rate of positivity for 19F higher than reports from other carriage studies in recent years. While both of these serotypes are targeted by PCV13 (the only conjugate vaccine recommended in the US through the period of the study), these serotypes have persisted as causes of IPD.^30^

The prevalence and dynamics of pneumococcal carriage in healthcare settings should be an important consideration for community transmission, particularly to at-risk individuals. Our study demonstrated a notable prevalence of pneumococcal carriage among healthcare workers, extending beyond those working in pediatric settings, and despite extensive use of non-pharmaceutical interventions during the study period. Together with the high frequency of detection of serotypes targeted by long-standing PCV13 immunization programs, this work contributes to the growing body of literature indicating that pneumococcal carriage, including vaccine-type pneumococci, has been underestimated in adults. Overall, our study highlights how improved understanding of the pneumococcus circulating in the community setting can explain the continued pneumococcal disease due to vaccine serotypes among adults and supports the direct vaccination of older adults.

## Data Availability

All data produced in the present study are available upon reasonable request to the authors

## Author’s contributions

ALW conceived the study. LRG, RAP, AA, BDG, DMW and ALW designed the study. MC, RD^2^, RAM, RAP and ALW managed the study. PW, NJV, MSH, DYC, LW, LC and MH were responsible for and performed the assays. PW, RD^1^, DMW and ALW performed the analyses and interpreted the data. PW, RD^1^ and ALW drafted the manuscript. All authors amended and commented on the final manuscript.

## Acknowledgements

We gratefully acknowledge the Yale IMPACT team for their efforts in supporting study participant recruitment, sample collection, and SARS-CoV-2 testing, and the participants for their time and commitment to the study.

